# Changes in Masking Policies in US Healthcare Facilities in the First Quarter of 2023: Do COVID-19 Cases, Hospitalizations, or Local Political Preferences Predict Loosening Restrictions?

**DOI:** 10.1101/2023.07.11.23292518

**Authors:** Sarah L. Miller, Vinay Prasad

## Abstract

**Introduction:** In the first half of 2023, many US healthcare facilities updated their policies on the required use of face masks by patients and employees. Despite statewide lifts of mask mandates in healthcare settings, the decisions by individual healthcare facilities have remained controversial and inconsistent. We sought to understand these decisions by examining COVID-19 case rates, hospitalization rates, and political affiliation, among counties where hospitals reported updated masking policies in news or social media posts.

**Methods:** We searched Twitter, Facebook, and Google for news stories related to changes in US healthcare facility masking policies between February 1st and April 30th, 2023. We extracted county-level COVID-19 cases and hospitalizations using data from the CDC and political affiliation was measured using the 2020 presidential election results. We performed logistic regression using COVID-19 cases, hospitalizations, and political affiliation as predictors and a complete lifting of masking requirements as the outcome.

**Results:** We found that the odds of lifting the mask requirement was not associated with COVID-19 cases (OR 1.00, 95% CI 0.97 - 1.02, p-value = 0.54), or hospitalizations (OR 1.06 95% CI 0.88-1.27, p-value = 0.33). We found that for every 10% increase in Republican votes in the 2020 presidential election, there was a 1.33 (95% CI 1.07 - 1.64, p-value = 0.01) increase in odds of having lifted masking requirements completely.

**Discussion:** We found that the odds of lifting the face mask requirement in healthcare facilities was not associated with COVID-19 cases or hospitalizations but was associated with county-level political affiliation. Our results raise the concern that public health measures may be increasingly seen as political gestures or a response to local political factors.

## Introduction

The lifting of statewide healthcare masking mandates in the first few months of 2023 resulted in many healthcare facilities updating their policies on patient and employee masking requirements. Though many dropped the requirement altogether, policy changes have been controversial, and some healthcare facilities have continued to require the use of masks. We sought to better understand these decisions by examining COVID-19 case rates, hospitalization rates, and political affiliation, in counties where healthcare facilities reported updated masking policies in news or social media posts.

## Methods

We searched Twitter, Facebook, and Google for news stories related to changes in US healthcare facility masking policies between February 1^st^ and April 30^th^, 2023. Our search terms included “mask” AND (“mandate” OR “hospital” OR “policy” OR “optional”). We recorded whether healthcare facilities completely lifted the mandate, required masks in certain units, or required masks in all clinical settings. COVID-19 cases per 100,000 and hospitalizations per 100,000 from February 1^st^ – April 30^th^ were recorded using county-level data from the CDC.^1^ Political affiliation was measured using 2020 county-level presidential election results^2^ and recorded as the proportion who voted for the Republican candidate. We performed logistic regression in Stata (version 17.0) using cases per 100,000, hospitalizations per 100,000, and political affiliation as predictors, and a complete lifting of mask mandates in the healthcare facility as the outcome.

## Results

We identified 316 hospitals from 36 states and 193 counties reporting masking policy updates in healthcare settings (**Supplemental Figure**). Of these, 66.1% (n = 209) did not have a masking requirement, 22.5% (n = 71) required masks in specific units, and 11.4% (n = 36) required masks in all clinical settings. We found that the odds of lifting the mask requirement was not associated with COVID-19 cases (OR 1.00, 95% CI 0.97 - 1.02, p-value = 0.54), or hospitalizations (OR 1.06 95% CI 0.88-1.27, p-value = 0.33). We found that for every 10% increase in Republican votes in the 2020 presidential election, there was a 1.33 (95% CI 1.07 - 1.64, p-value = 0.01) increase in odds of having lifted the masking requirement completely (**Figure)**.

**Figure.**
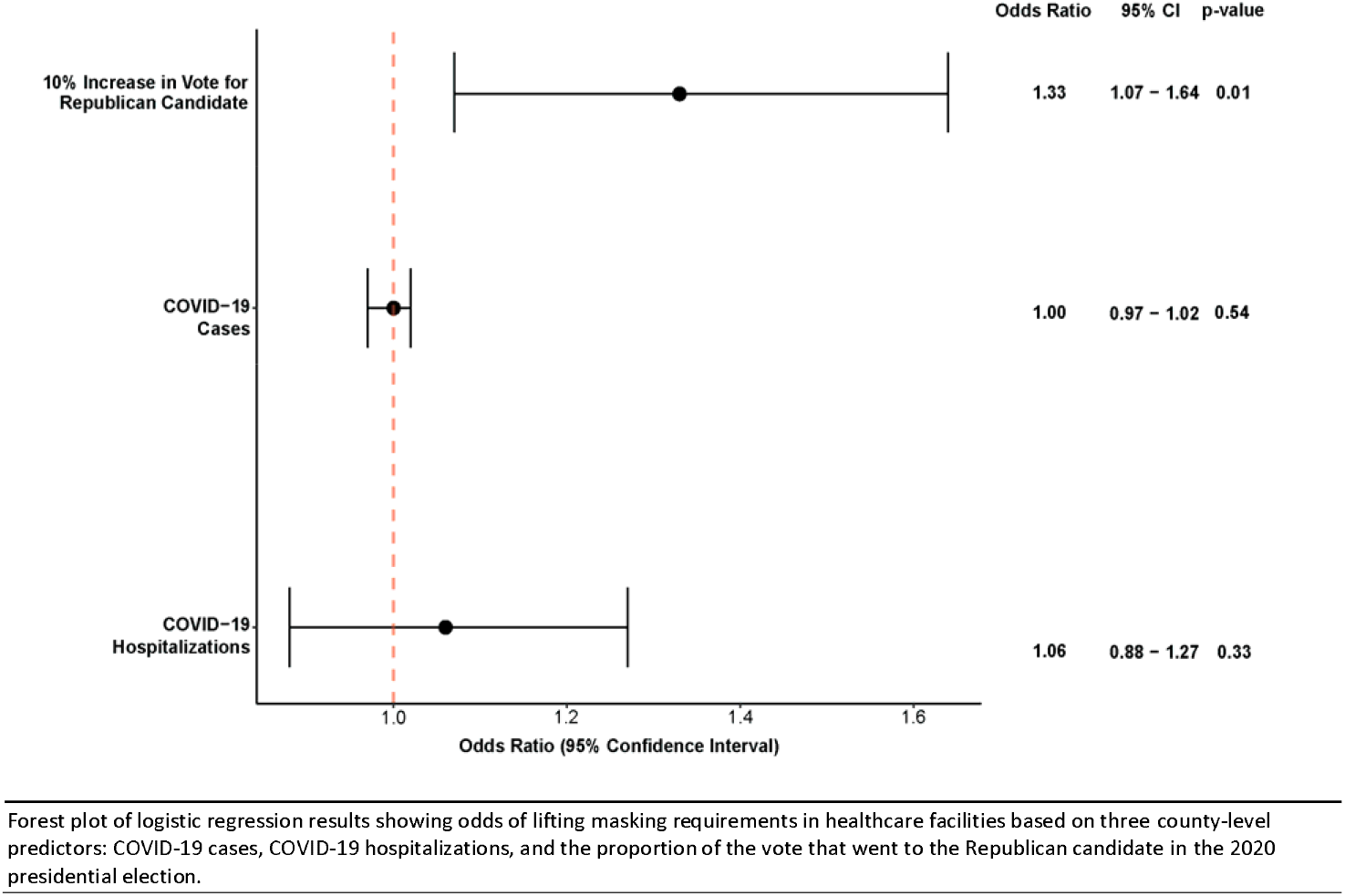
Odds of lifting masking requirement by county-level political affiliation, COVID-19 cases, and COVID-19 hospitalizations

## Discussion

Mask mandates in hospital settings has been a controversial topic,^3-5^ and over the early months of 2023, a number of hospitals relaxed or removed requirements. We found that these policy changes had no correlation with metrics of disease severity – cases or hospitalizations – but were correlated with the county level voting percentage for the Republican candidate based on 2020 presidential election results.

While we were only able to assess a limited number of healthcare facilities and counties based on news and social media posts, our results raise the concern that public health measures may be increasingly seen as political gestures or a response to local political factors.

## Data Availability

All data are publicly available

## Author Affiliations

Department of Epidemiology and Biostatistics, University of California, San Francisco

## Author Contributions

Miller had full access to all of the data in the study and takes responsibility for the integrity of the data and the accuracy of the data analysis.

## Concept and design

Prasad

## Acquisition, analysis, or interpretation of data

Miller, Prasad

## Drafting of the manuscript

Miller, Prasad

## Critical revision of the manuscript for important intellectual content

Miller, Prasad Statistical analysis: Miller

## Supervision

Prasad

## References

1. Centers for Disease Control and Prevention. COVID Data Tracker. Atlanta, GA: US Department of Health and Human Services, CDC; 2023, May 16. https://covid.cdc.gov/covid-data-tracker

2. Presidential election results: Live map of 2020 electoral votes. https://www.nbcnews.com. https://www.nbcnews.com/politics/2020-elections/president-results/

3. Shenoy, E. S., Babcock, H. M., Brust, K. B., Calderwood, M. S., Doron, S., Malani, A. N., Wright, S. B., & Branch-Elliman, W. (2023). Universal Masking in Health Care Settings: A Pandemic Strategy Whose Time Has Come and Gone, For Now. Ann Intern https://doi.org/10.7326/M23-0793

4. Palmore, T. N., & Henderson, D. K. (2023). For Patient Safety, It Is Not Time to Take Off Masks in Health Care Settings. Ann Intern Med. https://doi.org/10.7326/M23-1190

5. Prasad V. Hospital masking should be optional. Sensible Medicine. Published February 4, 2023. Accessed June 2, 2023. https://sensiblemed.substack.com/p/hospital-masking-should-be-optional

