## Supplemental Figure for "Changes in Masking Policies in US Healthcare Facilities in the First Quarter of 2023: Do COVID-19 Cases, Hospitalizations, or Local Political Preferences Predict Loosening Restrictions?"

**Supplemental Figure. Map of healthcare facility masking policy changes reported in news or social media posts between February 1^st^ – April 30^th^, 2023**

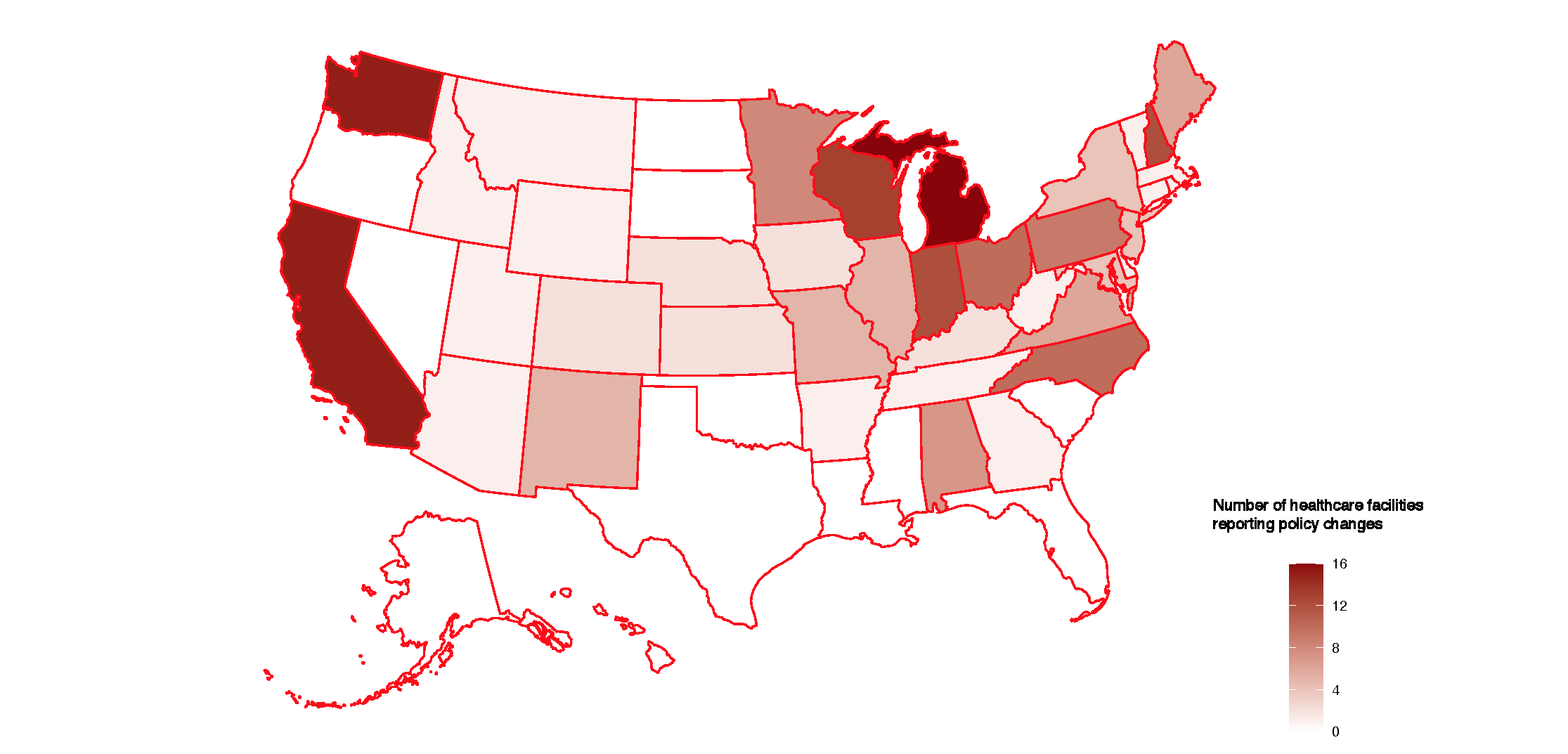

Number of healthcare facilities reporting policy changes
